# Assessment of Antibiotic Resistance and Associated Factors in Patients with Tuberculosis: A Nationwide Cross-Sectional Study of Indonesian TB-DRS

**DOI:** 10.1101/2025.04.16.25325965

**Authors:** Yuni Rukminiati, Felix Mesak, Dina Lolong, Pratiwi Sudarmono

## Abstract

**Objective:** Indonesia ranks second globally in terms of incidence of tuberculosis (TB), with a prevalence of 759 (95% CI: 589.7-960.8) cases per 100,000 people. Here, our cross-sectional population-based study (n=494) in Indonesia aims to reestablish the link between associated risk factors and antibiotic resistance.

**Method:** Data for this nationwide TB study was taken from the Tuberculosis Drug Resistance Survey (TB-DRS) by the Ministry of Health, Republic of Indonesia. The data was collected from a TB clinical field study conducted between 2017 and 2018. Participants provided demographic information through a self-report questionnaire after giving informed consent. Clinical samples were sent to our National Laboratory for testing the TB drug resistance status using MGIT960, GeneXpert, and Illumina-based WGS. Descriptive statistics were used to analyze the data with DATAtab software including Chi-square and other tests, and logistic regression. A significance level of p<0.05 was applied.

**Results:** TB resistance to drugs can be acquired irrespective of age-group (χ2=2.17, p=0.825), college education (OR: 1.1, 95% CI: 0.76-1.60, p=0.617, χ2=0.25), employment (OR: 0.93, 95% CI: 0.65-1.34, p=0.712, χ2=0.14), actively smoking (OR: 1.02, 95% CI: 0.61-1.69, p=0.951, χ2=0.00), geographical location (χ2=0.23, p=0.893), living community (OR: 0.94, 95% CI: 0.65-1.35, p=0.735, χ2=0.11), or previous attempts to seek treatment (OR: 1.26, 95% CI: 0.84-1.89, p=0.268, χ2=1.23). However, being female (OR: 1.6, 95% CI: 1.09-2.33, p=0.015, χ2=5.87), having a history of smoking (OR: 0.65, 95% CI: 0.45-0.94, p=0.021, χ2=5.36), and previously treated TB (OR: 0.02, 95% CI: 0.01-0.08, p<0.001, χ2=86.61) were linked to higher antibiotic resistance.

**Conclusion:** Focusing surveillance efforts on these statistically significant associated risk factors is crucial to halt the proliferation of drug-resistant TB within the population.

## Introduction

Tuberculosis (TB), caused by *Mycobacterium tuberculosis* (Mtb), remains a formidable global health challenge, particularly in densely populated areas, despite advancements in medicine including public healthcare management. An estimated 2 billion people, or 25% of the global population, has so far been infected with Mtb, underscoring the disease’s profound impact (Da Costa et al., 2024). Globally, TB continues to be a major source of morbidity and mortality. An estimated 1.6 million TB-related deaths occurred in 2021, with 187,000 of those deaths occurring in HIV-positive individuals (Da Costa et al., 2024). In high-burden countries, the morbidity rate of TB, a number of new and relapsed TB cases, can exceed 500 cases per 100,000 population. For example, South Africa reports a TB morbidity rate of around 615 cases per 100,000 people, whereas India has a morbidity rate of roughly 193 cases per 100,000 people (WHO, 2022). In 2021, the worldwide TB mortality rate was roughly 20 deaths per 100,000 individuals. Nevertheless, this figure is considerably elevated in countries with a high burden of TB. For instance, Nigeria’s TB mortality rate is approximately 67 per 100,000 population, whereas Indonesia has a rate of about 40 per 100,000 population (WHO, 2022). Yet, Indonesia ranks second globally in terms of incidence of TB, after India, with a prevalence of 759 (95% CI: 589.7-960.8) cases per 100,000 people (Noviyani et al., 2021).

Unfortunately, only 47% of the estimated cases of TB were reported or found in Indonesia (Lestari et al., 2022). Difficulties in finding new TB cases include socioeconomic variables such as underreporting, delayed diagnosis, and poor therapies that make managing diseases more difficult, Additionally, there is limited access to diagnostic facilities in many endemic places. As a result, a large number of TB cases are either misdiagnosed or discovered after the disease has spread to other people. The fact that many TB cases go unreported could also happen for a variety of reasons, including as lack of knowledge about the disease, fear of discrimination, and social shame. These are certainly another major obstacle. Another socioeconomic problem may include poverty, hunger, and substandard living conditions, which raise the risk of TB and make it more difficult to receive medical care. Crucially, the development of drug-resistant TB strains poses a significant obstacle to TB treatment. Also important to note that various elements lead to the emergence of drug-resistant TB, would be misuse of antibiotics and unfinished treatment regimens, in addition to insufficient access to quality medical care. Patients might not finish their treatment because of the expensive medication, prolonged therapy time, and adverse effects. Moreover, health care professionals might be deficient in the training and tools required to handle drug-resistant TB efficiently. Facing those drawbacks, however, Indonesia’s own TB statistics are still emerging quickly over time. There have been four times as many reported TB publications from Indonesia in only the last ten years from the Pubmed search (https://pubmed.ncbi.nlm.nih.gov/).

There are actually two main sources of concern with TB. (1) The emergence of TB treatment resistance is a concerning front in addition to (2) how to treat and prevent the spread of tuberculosis (Lestari et al., 2022). Thus far, every Mtb lineage discovered in Indonesia has developed treatment resistance, ranging from first-line antibiotic resistance to multidrug resistance TB (MDR) (Sinulingga et al., 2023). The predominant Mtb lineages evolving in Indonesia are L1, L2, and L4, with the L3 was being identified recently (Rukminiati et al., 2024). Yet, it is still essential to conduct population-based research to comprehend the relationship between patient characteristics, treatment outcomes, and drug resistance (Noviyani et al., 2021).

The objective of this study is to further update previously reported TB drug resistance and identify the associated societal epidemiological patterns that could support improved TB treatment and management in the archipelagic country. Based on a population-wide study of patients with TB conducted throughout Indonesia, we present the findings of a cross-sectional study evaluating antibiotic resistance and the associated factors.

## Methods

### Study design and setting

Data from Tuberculosis Drug Resistance Survey (TB-DRS) of the Ministry of Health, Republic of Indonesia were sampled for an assessment of a nationwide descriptive cross-sectional TB study reported here. The sampled data was retrieved from a TB clinical field study conducted between 2017 and 2018 and overseen by the Ministry of Health.

Indonesia is an archipelagic country in Southeast Asia at latitude 2°30′00.00″ South, longitude 117°54′00.00″ East. According to the TB-DRS record, clinical samples from TB patients were collected from various clinics and hospitals across the provinces of North, West, and South Sumatera, as well as Lampung on the island of Sumatera. In the island of Java, samples from TB patients were gathered from numerous clinics and hospitals in the provinces of West, Central, and East Java, along with Jakarta. Additionally, clinical samples from TB patients were sourced from a variety of clinics and hospitals in other islands located in Central and Eastern Indonesia, including West Kalimantan, North and South Sulawesi, Papua, and both Central and South Papua. Therefore, we evaluate the TB status from different areas of Indonesia during the specified timeframe, which will give us insights into both the country’s TB antibiotic resistance and its associated factors.

### Population

The study population comprised both male and female patients with TB at variety of age groups, i.e. 15-24, 25-34, 35-44, 45-54, 55-64, and 65+ years of age. Patients were classified based on their status of antibiotic susceptibility, treatment or ever treated, smoking or ever smoking, work, college education, place of living whether they were coming from urban or rural area, and the associated Mtb lineages. The inclusion criteria required patients to be TB positive as validated by MGIT960® cultured Mtb (Becton, Dickinson and Company, Franklin Lakes, NJ, USA), PCR-based GeneXpert® MTB/RIF Ultra (Cepheid, Sunnyvale, CA, USA) and the Illumina-based (Illumina Inc, San Diego, CA, USA) Mtb whole genome sequencing from the TB-DRS archives.

### Sample size and sampling technique

A total 494 patients with TB from TB-DRS were analyzed for the risk factors of antibiotic resistance and its associated factors. The sampling approach was conducted in adherence to the national guidelines, including the informed consent requirement, and supervised by the Ministry of Health. Consent form was formally signed by patients. All patients who were identified as having TB or drug-resistant TB received proper care. A maximum of 5 mL of sputum was collected from TB patients and dispatched to the Laboratory of Bacteriology at the National Laboratory of Prof. Sri Oemijati, Ministry of Health in Jakarta for microbiological and molecular testing. Both clinical samples and data generated from the tests were stored at the repository of TB-DRS. The clinical research was carried out in conformity with the principles of the Helsinki Declaration.

### Data collection

Per TB-DRS, data to be analyzed in this study were gathered using a self-administered questionnaire comprising ten sections: name, address, healthcare provider, age, sex, profession, education, treatment, past treatment, and smoking or ever smoking status. After obtaining ethical clearance from the Indonesian National Ethics Committee of the Ministry of Health, the data collection was conducted. In brief, all patients for TB screening were given the questionnaire to fill in. Patients’ confidentiality was protected by providing serial numbers on the questionnaire form. Data security was ensured by limiting access to the collected data and granting access only to the primary researchers for this study. Computerized data was password protected, physical data forms and patients samples were kept under lock in a secured facility.

### Data analysis

Data analysis was conducted using the DATAtab interactive statistical analysis (DATAtab Team, 2025). To describe sociodemographic factors and the levels of study variables, descriptive statistics such as percentages and frequencies were used. Statistical tests used: a chi-square test which is a statistical method employed to assess whether there is a noteworthy difference between the observed counts in categorical data and the counts that would be anticipated if the null hypothesis holds true. Then, two-sided Fisher Exact Test which is a statistical method utilized to identify if there is a significant association between two categorical variables, taking into account the possibility of the relationship going in either direction, and it is especially beneficial when working with small sample sizes where the conventional Chi-square test may be unreliable. Also, the Mann-Whitney U test which is designed to evaluate differences between two independent groups when the dependent variable is either ordinal or continuous but does not follow a normal distribution. Next, the Kruskal-Wallis test which is a non-parametric method for comparing medians of three or more independent groups, particularly with non-normally distributed data or unequal variances, and is an extension of the Mann-Whitney U test for multiple groups. On the other hand, the Dunn-Bonferroni test identifies which pairs of groups show significant differences after a Kruskal-Wallis test, adjusting p-values using the Bonferroni correction to minimize Type I error from multiple comparisons. Multivariable logistic regressions were used to investigate the relationship between the independent factors and the result variable after the assumptions were confirmed. To find independent predictors of satisfaction, the β coefficient (adjusted odds ratio) was employed. A p-value of less than 0.05 indicated that a variable was statistically significant. Binary traits were used to construct the phylogenetic tree of the TB lineages, and the binary matrix was produced in PHYLIP file format (Felsenstein, 1989). Using IQ-TREE, a maximum-likelihood phylogenies was generated with binary data as the input (Nguyen et al., 2015). Binary sequences that were identical were maintained during alignment. In order to root the tree, an outgroup taxon name employs the sole Mtb L3 (ID #S69070) discovered in the TB-DRS database. The bootstrap replicate count was fixed at 1000. The Microreact pipeline was utilized to visualize the lineage tree and its correlation with risk factors for tuberculosis (Argimón et al., 2016).

### Ethics approval

The Indonesian National Ethics Committee Ministry of Health, Health Institute for Research and Development (*Komisi Etik Litbangkes Kemenkes RI*) gave its approval to the current study.

## Results

### Sociodemographic profile of patients

Out of 494 TB patients examined in this research, the majority were from two islands: Sumatera with 154 individuals (31.17%) and Java with 212 individuals (42.91%) (Fig. 1, Table 1). The remaining patients from Central-East Indonesia (CEI), totaling 128 (25.91%), were from the islands of Borneo (Kalimantan), Celebes (Sulawesi), and Papua (Fig. 1, Table 1). The Mtb isolated from the patients’ samples displayed a drug resistance phenotype of 38% for both the Sumatera and Java-Bali islands, while the CEI region exhibited a resistance rate of 41% (Fig. 1). Regarding the identified Mtb lineages, L4, L2, and L1 were the most prevalent in the country. L1 demonstrates a notable presence in the CEI at 30%, compared to 19% in Sumatera and 12% in Java-Bali (Fig. 1). In contrast, L2 exhibits a different pattern, with Sumatera hosting the majority of the lineages at 47%, while Java-Bali has 35% and CEI 25% (Fig. 1). Notably, the geographical distribution of L4 is the highest in Java-Bali at 52%, followed by CEI at 42% and Sumatera at 34% (Fig. 1).

**Figure 1.**
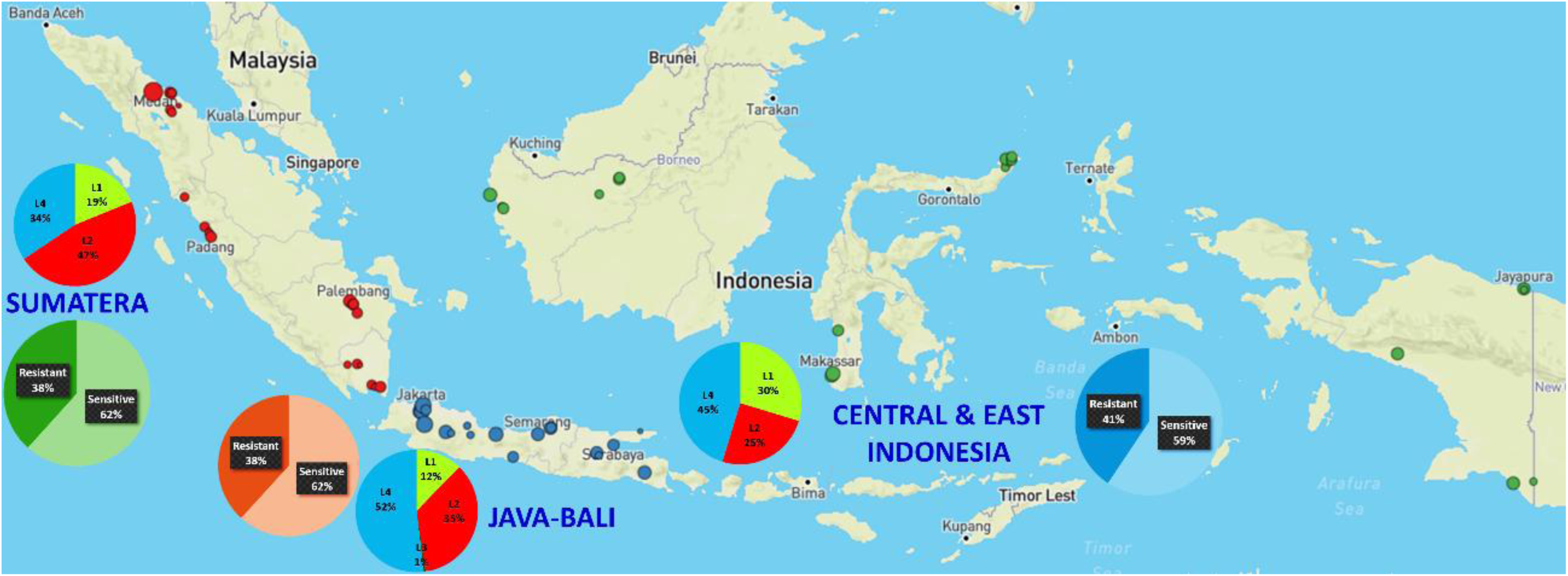
The location of patients participating in the study varied across the Indonesian archipelago. The highest percentage of TB patients was recorded in Java (42.91%) and Sumatera (31.17%), while the remaining patients were located in Central-East Indonesia (CEI, 25.91%). The percentages of Mtb drug resistance and the different lineages identified in these three Indonesian regions are provided.

**Table 1.**
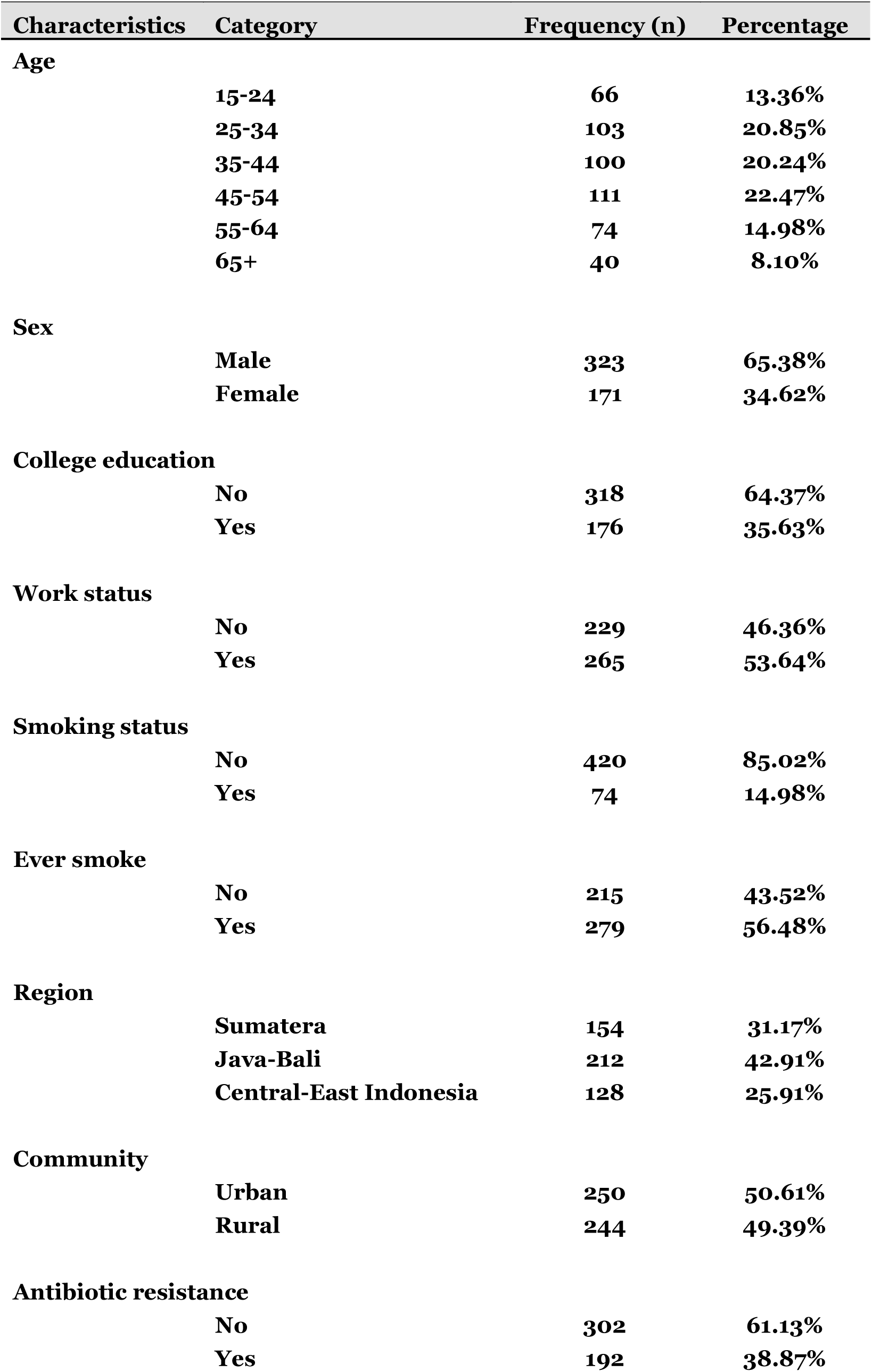
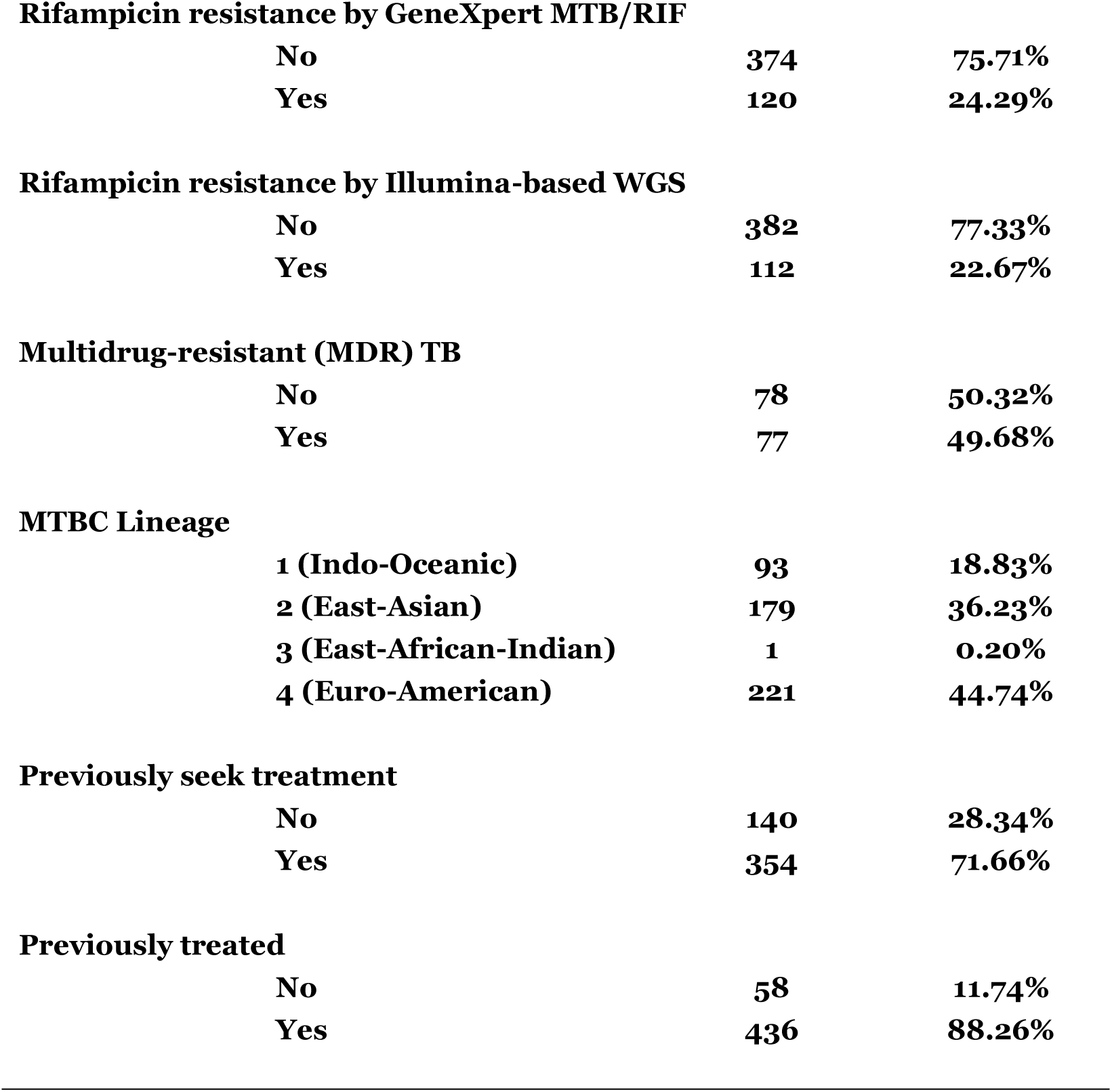
Sociodemographic characteristics of the TB patients (n=494)

| Characteristics | Category | Frequency (n) | Percentage |
| --- | --- | --- | --- |
| **Age** |  |  |  |
|  | 15-24 | 66 | 13.36% |
|  | 25-34 | 103 | 20.85% |
|  | 35-44 | 100 | 20.24% |
|  | 45-54 | 111 | 22.47% |
|  | 55-64 | 74 | 14.98% |
|  | 65+ | 40 | 8.10% |
| **Sex** |  |  |  |
|  | Male | 323 | 65.38% |
|  | Female | 171 | 34.62% |
| **College education** |  |  |  |
|  | No | 318 | 64.37% |
|  | Yes | 176 | 35.63% |
| **Work status** |  |  |  |
|  | No | 229 | 46.36% |
|  | Yes | 265 | 53.64% |
| **Smoking status** |  |  |  |
|  | No | 420 | 85.02% |
|  | Yes | 74 | 14.98% |
| **Ever smoke** |  |  |  |
|  | No | 215 | 43.52% |
|  | Yes | 279 | 56.48% |
| **Region** |  |  |  |
|  | Sumatera | 154 | 31.17% |
|  | Java-Bali | 212 | 42.91% |
|  | Central-East Indonesia | 128 | 25.91% |
| **Community** |  |  |  |
|  | Urban | 250 | 50.61% |
|  | Rural | 244 | 49.39% |
| **Antibiotic resistance** |  |  |  |
|  | No | 302 | 61.13% |
|  | Yes | 192 | 38.87% |

**Rifampicin resistance by GeneXpert MTB/RIF**
|  |  |  |
| --- | --- | --- |
| No | 374 | 75.71% |
| Yes | 120 | 24.29% |

**Rifampicin resistance by Illumina-based WGS**
|  |  |  |
| --- | --- | --- |
| No | 382 | 77.33% |
| Yes | 112 | 22.67% |

**Multidrug-resistant (MDR) TB**
|  |  |  |
| --- | --- | --- |
| No | 78 | 50.32% |
| Yes | 77 | 49.68% |

**MTBC Lineage**
|  |  |  |
| --- | --- | --- |
| 1 (Indo-Oceanic) | 93 | 18.83% |
| 2 (East-Asian) | 179 | 36.23% |
| 3 (East-African-Indian) | 1 | 0.20% |
| 4 (Euro-American) | 221 | 44.74% |

**Previously seek treatment**
|  |  |  |
| --- | --- | --- |
| No | 140 | 28.34% |
| Yes | 354 | 71.66% |

**Previously treated**
|  |  |  |
| --- | --- | --- |
| No | 58 | 11.74% |
| Yes | 436 | 88.26% |

Recruited TB patients for the study were 323 men (65.38%) and 171 women (34.62%). A similar proportion of patients (50.61% and 49.39%, respectively) were recruited from urban and rural areas. Patients were divided into 6 age groups, with the following age distributions: 13.36% for those in the 15–24 age range, 20.85% for those in the 25–34 age range, 20.24% for those in the 35–44 age range, 22.47% for those in the 45–54 age range, 14.98% for those in the 55–64 age range, and 8.10% for those over 65. Of those patients, 265 (53.64%) had a job, whereas the remaining 46.36% did not. The majority of the patients (64.37%) had either completed high school, middle school, or elementary school, while 176 patients (35.63%) had completed college education. Despite the decrease in smokers, 14.98% of the patients, or 74, continued to smoke. Of those who have ever smoked, 56.48% have previously, even though some had already quit. We discovered that 192 patients (38.87%) were resistant to TB treatment, with 120 (24.29%) or 112 (22.67%) resistant to the first-line TB drug Rifampicin by Xpert® or Illumina-based WGS, respectively. It is worth noting that the two methodologies used had 1.62% disparities in Rifampicin detection. Of the individuals who have drug-resistant TB, 77 had multi-antibiotic-resistant Mtb. In fact, 71.66% of patients sought treatment, and 88.26% of them received it.

### Associated factors toward antibiotic resistance among TB patients

A summary of associated factors toward antibiotic resistance among TB patients identified in our populational study is given in Table 2.

**Table 2.**
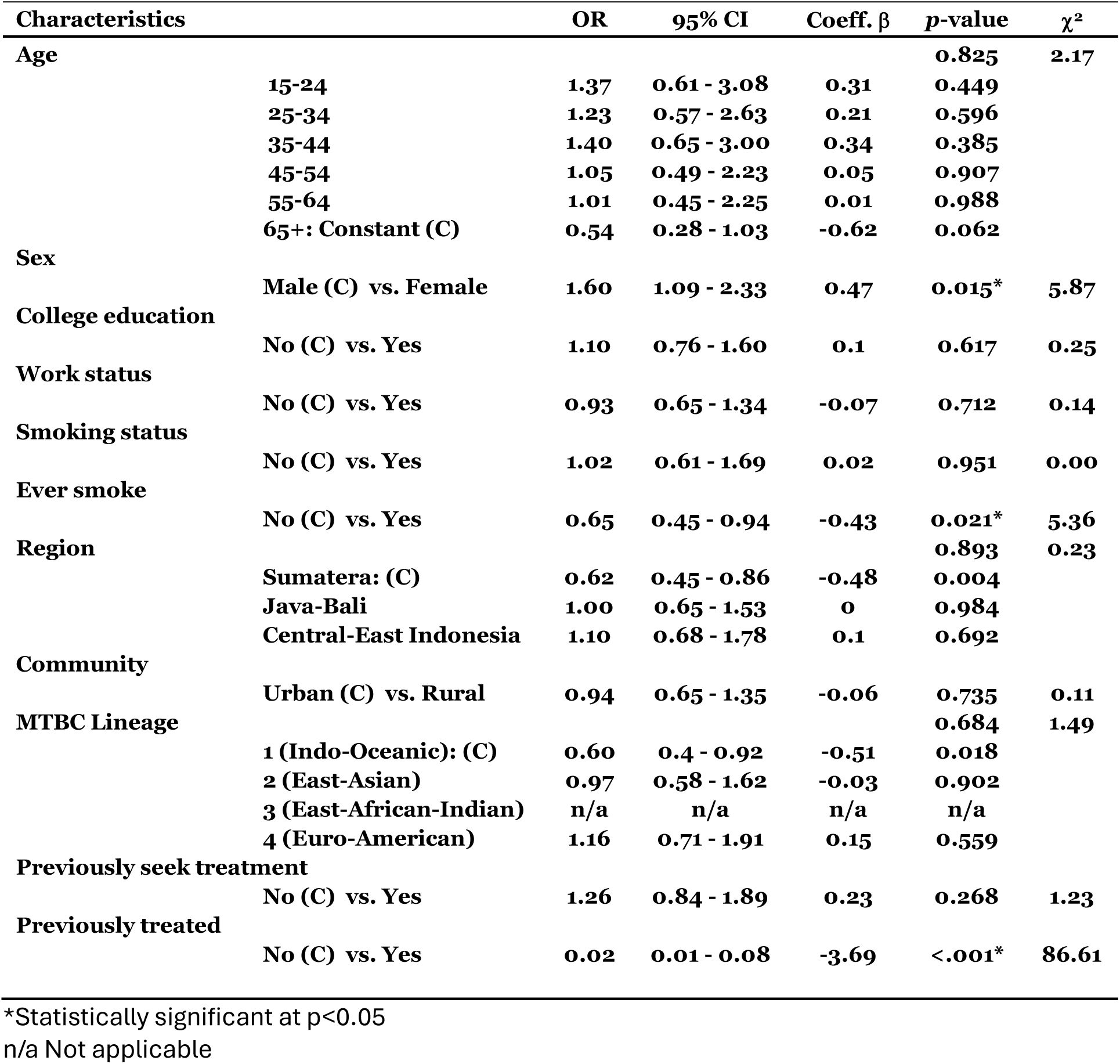
Results from the logistic regression model.

| Characteristics | | OR | 95% CI | Coeff. $\beta$ | p-value | $\chi^2$ |
| --- | --- | --- | --- | --- | --- | --- |
| **Age** |  |  |  |  | **0.825** | **2.17** |
|  | 15-24 | 1.37 | 0.61 - 3.08 | 0.31 | 0.449 |  |
|  | 25-34 | 1.23 | 0.57 - 2.63 | 0.21 | 0.596 |  |
|  | 35-44 | 1.40 | 0.65 - 3.00 | 0.34 | 0.385 |  |
|  | 45-54 | 1.05 | 0.49 - 2.23 | 0.05 | 0.907 |  |
|  | 55-64 | 1.01 | 0.45 - 2.25 | 0.01 | 0.988 |  |
|  | 65+: Constant (C) | 0.54 | 0.28 - 1.03 | -0.62 | 0.062 |  |
| **Sex** |  |  |  |  |  |  |
|  | Male (C) vs. Female | 1.60 | 1.09 - 2.33 | 0.47 | 0.015* | 5.87 |
| **College education** |  |  |  |  |  |  |
|  | No (C) vs. Yes | 1.10 | 0.76 - 1.60 | 0.1 | 0.617 | 0.25 |
| **Work status** |  |  |  |  |  |  |
|  | No (C) vs. Yes | 0.93 | 0.65 - 1.34 | -0.07 | 0.712 | 0.14 |
| **Smoking status** |  |  |  |  |  |  |
|  | No (C) vs. Yes | 1.02 | 0.61 - 1.69 | 0.02 | 0.951 | 0.00 |
| **Ever smoke** |  |  |  |  |  |  |
|  | No (C) vs. Yes | 0.65 | 0.45 - 0.94 | -0.43 | 0.021* | 5.36 |
| **Region** |  |  |  |  | **0.893** | **0.23** |
|  | Sumatera: (C) | 0.62 | 0.45 - 0.86 | -0.48 | 0.004 |  |
|  | Java-Bali | 1.00 | 0.65 - 1.53 | 0 | 0.984 |  |
|  | Central-East Indonesia | 1.10 | 0.68 - 1.78 | 0.1 | 0.692 |  |
| **Community** |  |  |  |  |  |  |
|  | Urban (C) vs. Rural | 0.94 | 0.65 - 1.35 | -0.06 | 0.735 | 0.11 |
| **MTBC Lineage** |  |  |  |  | **0.684** | **1.49** |
|  | 1 (Indo-Oceanic): (C) | 0.60 | 0.4 - 0.92 | -0.51 | 0.018 |  |
|  | 2 (East-Asian) | 0.97 | 0.58 - 1.62 | -0.03 | 0.902 |  |
|  | 3 (East-African-Indian) | n/a | n/a | n/a | n/a |  |
|  | 4 (Euro-American) | 1.16 | 0.71 - 1.91 | 0.15 | 0.559 |  |
| **Previously seek treatment** |  |  |  |  |  |  |
|  | No (C) vs. Yes | 1.26 | 0.84 - 1.89 | 0.23 | 0.268 | 1.23 |
| **Previously treated** |  |  |  |  |  |  |
|  | No (C) vs. Yes | 0.02 | 0.01 - 0.08 | -3.69 | <.001* | 86.61 |

\*Statistically significant at $p < 0.05$
n/a Not applicable

In the age group, logistic regression study was conducted to predict the value Mtb “*Resistant*” by analysing the effects of age groups 15–24, 25–34, 35–44, 45–54, 55–64, and 65+ as a constant (C) or reference on variable antibiotic resistance. The model is not significant as a whole, according to the analysis. (χ^2^(5) = 2.17, p-value 0.825, n = 494). For every age group under or equal to 65, the coefficients of variables (β) are all positive (0.31, 0.21, 0.34, 0.05, and 0.01, respectively), increasing the likelihood that the dependent variable will be “*Resistant*”. All of the age-group’s p-values (0.449, 0.596, 0.385, 0.907, and 0.988, in that order) do not, however, show statistical significance. It is interesting to note that the likelihood that the dependent variable is “*Resistant*” increases by 1.37 (for 15–24), 1.23 (for 25–34), 1.40 (for 35–44), 1.05 (for 45–54), and 1.01 (for 55–64) times if the age group is the variable.

Among the 323 males and 171 females with TB, 113 (34.99%) and 79 (46.20%), respectively, were resistant to TB medications. The hypotheses test asked if the two categorical variables were independent of one another? The significance level for this study is set at 0.05. Antibiotic resistance and sex were compared using a Chi-square test. The Chi-square test satisfied its assumptions as all expected cell frequencies exceeded 5. A statistically significant correlation was found between antibiotic resistance and sex, χ²(1) = 5.92, p = 0.015, Cramér’s V = 0.11. An additional two-sided Fisher exact test was run comparing the two categories. Antibiotic resistance was significantly associated with sex (p = 0.016). The estimated p-value of 0.015 falls below the 5% threshold for significance. The Chi-square is therefore significant, and the null hypothesis is rejected. The impact of the variable antibiotic resistance on the female sex in order to predict the value “*Resistant*” was investigated using logistic regression analysis. A logistic regression analysis demonstrates the significance of the model overall (χ^2^(1) = 5.87, p = 0.015, n = 494). The female variable’s coefficient, β = 0.47, indicates a positive value. This indicates that the likelihood of the dependent variable being “*Resistant*” rises if the variable’s value is sex (female). The statistical significance of this influence is indicated by the p-value of 0.015. With an odds ratio of 1.6, there is a 1.6-fold increase in the likelihood that the dependent variable is “*Resistant*” if the variable is female sex.

Among patients with a college degree, 71 patients (40.34%) showed resistance to TB treatment. Comparably, 38.05% of persons with less education than a college degree fell into this category. College degree and antibiotic resistance did not significantly correlate, according to statistical analysis χ²(1) = 0.25, p = 0.617, Cramér’s V = 0.02. At 0.617, the computed p-value is higher than the predetermined significance limit of 5%. The Chi-square test is not significant; hence the null hypothesis is not rejected. Logistic regression analysis was performed to examine the influence of college education on variable antibiotic resistance to predict the value “*Resistant*”. The model is not significant as a whole, according to logistic regression analysis (χ^2^(1) = 0.25, p = 0.617, n = 494). The college education variable has a positive coefficient (β = 0.1). This suggests that if the variable’s value is a college education, there is a higher likelihood that the dependent variable will be “*Resistant*”. The p-value of 0.617, however, suggests that there is no statistical significance for this influence. The dependent variable’s likelihood of being “*Resistant*” increases by 1.1 times if the independent variable is college education, according to the odds ratio of 1.1.

Patients who were employed but resistant to TB treatment accounted for 38.11%. Comparably, it was discovered that 39.74% of patients without jobs were resistant to TB medication. There was no statistically significant correlation found between the work status and antibiotic resistance, χ²(1) = 0.14, p = 0.712, Cramér’s V = 0.02. The model as a whole is consequently not significant, and the null hypothesis is not rejected. The employed patients’ coefficient of the variable (β) is -0.07, indicating a negative value. This indicates that the likelihood of the dependent variable being “*Resistant*” falls if the variable’s value is being used. With an odds ratio of 0.93, there is a 0.93-fold increase in the likelihood that the dependent variable is “*Resistant*” if the variable is used.

Patients who were currently smoking or had ever smoked had developed resistance to TB medications in 39.19% and 34.41% of cases, respectively. However, it was discovered that there was a statistically conflicting significant association between the two smoking statuses (smoker: χ²(1)=0, p = 0.951, Cramér’s V = 0, ever smoked: χ²(1)=5.36, p = 0.021, Cramér’s V = 0.1) and antibiotic resistance based on the results of the Chi-square and Fisher exact test. As a result, whereas the null hypothesis is rejected for individuals who had ever smoked, it is not rejected for patients who smoke currently. The coefficients of the variable status, β=0.02 and β=-0.43, respectively, explained the difference between those smokers and the patients who had ever smoked. It implies that the likelihood that the dependent variable will be “*Resistant*” rises if the variable’s value smokes and falls if the variable’s value has ever smoked.

The vast archipelagic nation of Indonesia may have an impact on the distribution of patients who are resistant to TB treatment. The proportion of patients resistant to TB antibiotic therapy was determined to be between 38 and 40% in three different regions of Indonesia: Sumatera, Java-Bali, and Central-East Indonesia. However, our research indicates that there was no statistically significant correlation found between the area and antibiotic resistance, χ²(2) = 0.23, p = 0.893, Cramér’s V = 0.02. Thus, the null hypothesis is not rejected.

In this investigation, it is being determined whether patients’ living arrangements—rural or urban—may have an impact on the emergence of TB treatment resistance. Undoubtedly, there was no discernible difference in the likelihood of antibiotic resistance development between patients residing in rural (38.11%) and urban (39.60%) areas. Antibiotic resistance and the patients’ communities did not statistically significantly correlate, χ²(1) = 0.11, p = 0.735, Cramér’s V = 0.02. Thus, the null hypothesis is not rejected.

The possibility that one Mtb lineage may develop TB antibiotic resistance more than the other is also being investigated. Our null hypothesis, then, is that there is no difference between the dependent variable (Mtb lineages) and antibiotic resistance. The difference between antibiotic resistance and the dependent variable Mtb lineages was not statistically significant for the presented data, according to a Mann-Whitney U-Test, U = 27845, n1 = 192, n2 = 302 p = 0.459. Thus, the null hypothesis is not rejected. A logistic regression analysis was conducted to predict the value “*Resistant*” by analysing the impact of Mtb L4, L2, and L3 on the variable antibiotic resistance. According to logistic regression analysis, the model is not significant overall (χ^2^(3) = 1.84, p = 0.607, n = 494). The variable Mtb L4 has a positive coefficient of b = 0.15. This indicates that there is a greater chance that the dependent variable will be “*Resistant*” if the variable’s value is Mtb L4. The p-value of 0.559, however, suggests that there is no statistical significance for this influence. With an odds ratio of 1.16, there is a 1.16-fold increase in the likelihood that the dependent variable is “*Resistant*” if the variable is Mtb L4. The coefficient of the variable Mtb L2 is b = -0.03, which is negative, and it points in the slightly opposite direction. The influence is not statistically significant, though, as indicated by the p-value of 0.902.

There were two results, depending on whether patients sought treatment or were actually receiving antibiotics. However, it was discovered that there was a statistically conflicting significant association between the two treatment statuses (previously seeking treatment: χ²(1) = 1.23, p = 0.268, Cramér’s V = 0.05, previously treated: χ²(1) = 86.61, p = <.001, Cramér’s V = 0.42) and antibiotic resistance based on the results of the Chi-square and Fisher exact test. As a result, whereas the null hypothesis is rejected for individuals who had been treated with antibiotic, it is not rejected for patients who sought for TB treatment. For patients who sought therapy and had antibiotic resistance, the variable’s coefficient is β = 0.23, which is positive. This indicates that there is a higher chance that the dependent variable will be “*Resistant*” if the variable’s value is patients who sought treatment. Nevertheless, the statistical significance of this influence is not shown by the p-value of 0.268. With a 1.26 odds ratio, there is a 1.26-fold increase in the likelihood that the dependent variable is “*Resistant*” if the variable is patients who sought treatment. In contrast, the coefficient of the variable for patients who were treated and developed a resistance to the TB treatment is β = - 3.69, indicating a negative correlation. This implies that the likelihood of the dependent variable being “*Resistant*” falls if the value of the variable is patients who have received treatment. The statistical significance of this influence is indicated by the p-value of <0.001.

A summary of odd ratios for Mtb lineages and the isolated regions found in our population study are provided in Table 3. Our null hypothesis states that the dependent variable, Mtb lineage, is the same in all three separate locations (Sumatera, Java-Bali, and Central-East Indonesia). A Kruskal-Wallis test demonstrated that there is a significant difference between the categories of the independent variable with regard to the dependent variable Mtb lineages, p=0.002. Thus, with the available data, the null hypothesis is rejected. According to the results of the Dunn-Bonferroni test, the pairwise group comparisons between Java-Bali-Central-East Indonesia and Sumatera-Java-Bali have an adjusted p-value 0.035 and 0.004, respectively. As a result, it is reasonable to assume that these groups are significantly different from one another based on the available data. The impact of Mtb L4 (Euro-American), Mtb L2 (East Asian), and Mtb L3 (East African-Indian) on the variable region to predict: (1) the value “*Sumatera*”, (2) the value “*Java-Bali*”, and (3) “*Central-East Indonesia*” were investigated using logistic regression analysis. Logistic regression analysis shows that the model as a whole is significant for “*Sumatera*” (χ^2^(3) = 12.87, p = 0.005, n = 494), “*Java-Bali*” (χ^2^(3) = 14.87, p = 0.002, n = 494) and “*Central-East Indonesia*” (χ^2^(3) = 16.95, p = 0.001, n = 494).

**Table 3.** Results from the logistic regression for important Mtb lineage factors in a specific Indonesian region using simple logistic regression.

| Variable | Category | OR | 95% CI | Coeff. $\beta$ | p-value | $\chi^2$ |
| --- | --- | --- | --- | --- | --- | --- |
| **Region:<sup>c</sup>** |  |  |  |  | **0.002\*** | **12** |
| **Sumatera** | **1 (Indo-Oceanic): (C)** | **0.45** | **0.29 - 0.7** | **-0.79** | **<.001\*** |  |
|  | **2 (East-Asian)** | **1.49** | **0.87 - 2.52** | **0.4** | **0.144** |  |
|  | **3 (East-African-Indian)** | **n/a** | **n/a** | **n/a** | **n/a** |  |
|  | **4 (Euro-American)** | **0.70** | **0.41 - 1.19** | **-0.4** | **0.186** |  |
| **Java-Bali** | **1 (Indo-Oceanic): (C)** | **0.39** | **0.25 - 0.61** | **-0.95** | **<.001\*** |  |
|  | **2 (East-Asian)** | **1.86** | **1.08 - 3.19** | **0.6** | **0.025\*** |  |
|  | **3 (East-African-Indian)** | **n/a** | **n/a** | **n/a** | **n/a** |  |
|  | **4 (Euro-American)** | **2.55** | **1.51 - 4.31** | **0.9** | **<.001\*** |  |
| **Central-East Indonesia** | **1 (Indo-Oceanic): (C)** | **0.69** | **0.46 - 1.04** | **-0.37** | **0.08** |  |
|  | **2 (East-Asian)** | **0.32** | **0.18 - 0.55** | **-1.2** | **<.001\*** |  |
|  | **3 (East-African-Indian)** | **n/a** | **n/a** | **n/a** | **n/a** |  |
|  | **4 (Euro-American)** | **0.52** | **0.31 - 0.86** | **-0.7** | **0.011\*** |  |

<sup>c</sup>Analyzed using Kruskal-Wallis Test
\*Statistically significant at p<0.05
n/a Not applicable

## Discussion

While age groups do not appear to have an impact on the probability of having an Mtb antibiotic resistance phenotype, the probability does increase for age groups 15–24, 25–34, and 35–44 by 37%, 23%, and 40%, respectively (Table 2). Younger age groups therefore have a propensity to harbor more drug-resistant Mtb than older generations (45+). When a normal person is younger, their immune system is at its peak, which might actively shield the lung tissue from damage by drawing in inflammatory cells and potentially developing tubercles, which would make TB treatment ineffective. It needs to be elucidated if the bacilli evolve a resistant phenotype as a result of the TB drug’s decreasing dosage entering and reaching the tubercle (Dheda et al., 2018). The majority of populational studies failed to find a relationship between the introduction date of the drug and the emergence date of TB drug resistance. This probably indicates that other factors, such the rate at which mutations occur spontaneously, the costs of TB drug resistance in vivo in terms of fitness, and the therapeutic combinations in which antibiotics have a tendency to be administered, influence the development of antibiotic resistance (Nimmo et al., 2022). In terms of TB drug resistance among male and female patients, our data revealed there is an indication of an association between sex and the risk factor. This contrasts with the results of a global survey, which in 86 out of 106 countries revealed no evidence of a link between sex and the risk of MDR/RR-TB in TB patients (McQuaid et al., 2020). However, the survey discovered that males have a higher chance of contracting MDR/RR-TB in countries with higher antibiotic resistance burdens (McQuaid et al., 2020). Even though there were only one-third of female patients in our study, their risk of acquiring antibiotic resistance was 60% higher than that of their male counterparts (Table 1, 2). Despite the stark differences between the two study results, they both emphasize the necessity of a sex-differentiated approach to TB case surveillance and treatment (McQuaid et al., 2020). The majority of risk factors in our study, such as patients’ college degree, employment status, smoking habit, history of treatment seeking, and the three distinct regions or community types where sampling was done, were found to be unrelated to the likelihood of TB drug resistance. Among the risk variables that may increase the chance of a high TB incidence is socioeconomic status, such as unemployment and low education (Imam et al., 2021). Our analysis, however, demonstrates that this was not the case in terms of their correlation with antibiotic resistance. Despite the general consensus that smoking cigarettes increases the likelihood of developing antibiotic resistance, our study indicates that the overall smoking habit in the Indonesian community has little bearing on the development of drug resistance (Feldman et al., 2024). For the subset of participants in our study who have ever smoked before, the correlation with TB drug resistance becomes significant. It implies that even after quitting, a smoker may still be predisposed to become resistant to antibiotics. The importance of tuberculosis therapeutic in controlling its spread cannot be overstated (Soedarsono et al., 2023). In our study, those who had previously sought treatment for tuberculosis were not predisposed to developing treatment resistance. TB patients who have previously received anti-TB medication for at least one month are considered previously treated cases. Our findings indicate that TB drug resistance is linked to patients who received treatment, indicating that the possibility of treatment failure may potentially play a role in the emergence of antibiotic resistance. The Mtb lineages are thought to have a role in the establishment of TB treatment resistance (McGrath et al., 2014). As of right now, Mtb has been divided into seven lineages and numerous sub-lineages, each of which has unique traits that have developed alongside the human populations in which they are found (Nimmo et al., 2022). Our earlier research indicates that L1, L2, and L4 made up the majority of the lineages found in the Indonesian population, with L3 emerging as a new member (Rukminiati et al., 2024). We do not discover any correlation between Mtb lineages and medication resistance to TB, nevertheless. Upon constructing a phylogenetic binary tree using lineages and sub-lineages identified from patients’ Mtb samples, we found widespread antibiotic-resistant Mtb in all lineages present in the population (Figure 2). It is known that compared to other Mtb lineages, the four identified in Indonesia have a greater mutational rate (Jones et al., 2022). The dynamic of Mtb lineages distribution is evident when we divide patients from various Indonesian regions into three categories: Sumatera, Java-Bali, and Central-East Indonesia (Table 3). Certain lineages (L2, L4) were associated significantly to the Java-Bali and Central-East Indonesia regions. It is unclear whether there was a higher tendency for some lineages inside the subpopulation to develop than for others, or if only the lineages that initially enter the subpopulation may evolve within it (Nimmo et al., 2022).

**Figure 2.**
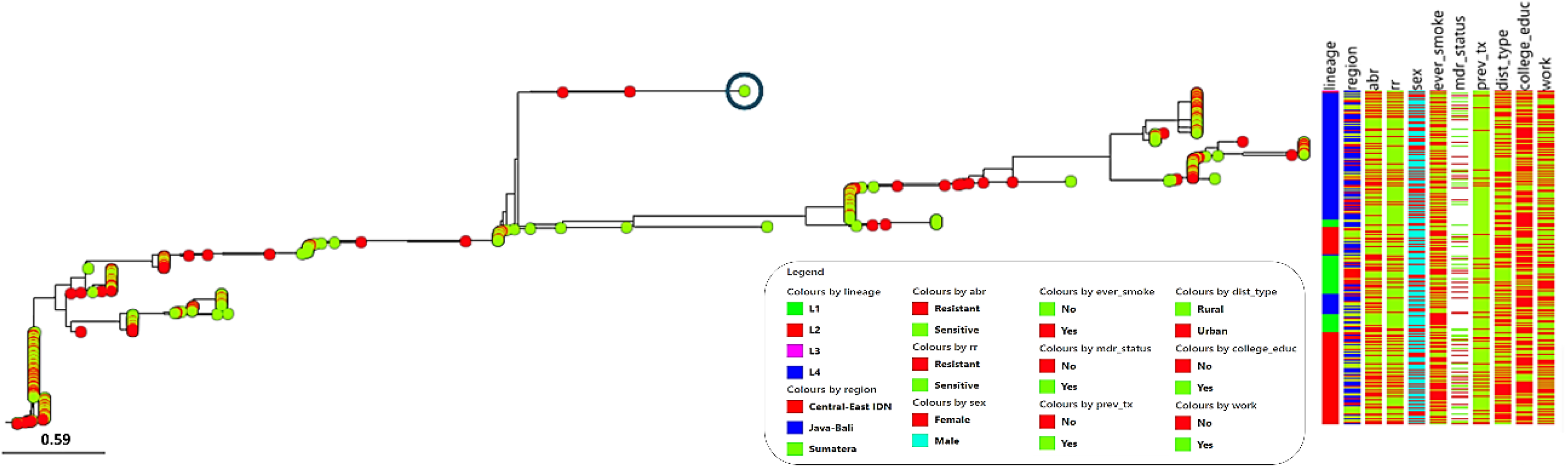
The TB lineages tree, which is rooted in the sole L3 (circled), is linked to associated risk factors for TB drug resistance (red dots: resistant, green dots: sensitive). Abbreviations: abr, antibiotic resistance; rr, (first-line antibiotic) rifampicin resistance, mdr, multi-drug resistance, prev_tx, previously treated, dist_type, type of district.

## Conclusion

With the exception of being female, having ever smoked, and having received prior antibiotic treatment, the majority of risk factors are unrelated to the likelihood of developing TB antibiotic resistance, according to our population-wide study conducted throughout Indonesia. Antibiotic resistance can develop in all three of the major Mtb lineages found in Indonesia, L1, L2, and L4, irrespective of the associated risk factors that the population may present. Three separate patient sample regions demonstrate a dynamic proportion of different Mtb lineages, indicating which lineage was either the first to enter the local population or expanded more quickly than others. Therefore, it is critical to treat TB patients as soon as possible while concentrating on the specific associated risk factors found in this study, irrespective of Mtb lineages.

## Data Availability

All data produced in the present work are contained in the manuscript

## Acknowledgments

The authors would like to thank everyone at NIHRD, Ministry of Health of Indonesia, LMK FKUI, Laboratorium Terpadu FKUI, HUMRC Makassar, BBLK Makassar, and BBLK Surabaya who supplied samples and clinical data to this study.

## References

Argimón, S., Abudahab, K., Goater, R. J. E., Fedosejev, A., Bhai, J., Glasner, C., Feil, E. J., Holden, M. T. G., Yeats, C. A., Grundmann, H., Spratt, B. G., & Aanensen, D. M. (2016). Microreact: Visualizing and sharing data for genomic epidemiology and phylogeography. Microbial Genomics, 2(11), e000093. 10.1099/mgen.0.000093

Da Costa, C., Benn, C. S., Nyirenda, T., Mpabalwani, E., Grewal, H. M. S., Ahmed, R., Kapata, N., Nyasulu, P. S., Maeurer, M., Hui, D. S., Goletti, D., & Zumla, A. (2024). Perspectives on development and advancement of new tuberculosis vaccines. International Journal of Infectious Diseases, 141, 106987. 10.1016/j.ijid.2024.106987

DATAtab Team. (2025). DATAtab: Online Statistics Calculator. [Computer software]. DATAtab e.U. https://datatab.net

Dheda, K., Lenders, L., Magombedze, G., Srivastava, S., Raj, P., Arning, E., Ashcraft, P., Bottiglieri, T., Wainwright, H., Pennel, T., Linegar, A., Moodley, L., Pooran, A., Pasipanodya, J. G., Sirgel, F. A., van Helden, P. D., Wakeland, E., Warren, R. M., & Gumbo, T. (2018). Drug-Penetration Gradients Associated with Acquired Drug Resistance in Patients with Tuberculosis. American Journal of Respiratory and Critical Care Medicine, 198(9), 1208–1219. 10.1164/rccm.201711-2333OC

Feldman, C., Theron, A. J., Cholo, M. C., & Anderson, R. (2024). Cigarette Smoking as a Risk Factor for Tuberculosis in Adults: Epidemiology and Aspects of Disease Pathogenesis. Pathogens, 13(2), 151. 10.3390/pathogens13020151

Felsenstein, J. (1989). PHYLIP - Phylogeny Inference Package (Version 3.2). Cladistics, 5, 164–166.

Imam, F., Sharma, M., Obaid Al-Harbi, N., Rashid Khan, M., Qamar, W., Iqbal, M., Daud Ali, M., Ali, N., & Khalid Anwar, M. (2021). The possible impact of socioeconomic, income, and educational status on adverse effects of drug and their therapeutic episodes in patients targeted with a combination of tuberculosis interventions. Saudi Journal of Biological Sciences, 28(4), 2041–2048. 10.1016/j.sjbs.2021.02.004

Jones, R. M., Adams, K. N., Eldesouky, H. E., & Sherman, D. R. (2022). The evolving biology of Mycobacterium tuberculosis drug resistance. Frontiers in Cellular and Infection Microbiology, 12, 1027394. 10.3389/fcimb.2022.1027394

Lestari, T., Kamaludin, null, Lowbridge, C., Kenangalem, E., Poespoprodjo, J. R., Graham, S. M., & Ralph, A. P. (2022). Impacts of tuberculosis services strengthening and the COVID-19 pandemic on case detection and treatment outcomes in Mimika District, Papua, Indonesia: 2014-2021. PLOS Global Public Health, 2(9), e0001114. 10.1371/journal.pgph.0001114

McGrath, M., Gey Van Pittius, N. C., Van Helden, P. D., Warren, R. M., & Warner, D. F. (2014). Mutation rate and the emergence of drug resistance in Mycobacterium tuberculosis. Journal of Antimicrobial Chemotherapy, 69(2), 292–302. 10.1093/jac/dkt364

McQuaid, C. F., Horton, K. C., Dean, A. S., Knight, G. M., & White, R. G. (2020). The risk of multidrug- or rifampicin-resistance in males *versus* females with tuberculosis. European Respiratory Journal, 56(3), 2000626. 10.1183/13993003.00626-2020

Nguyen, L.-T., Schmidt, H. A., Von Haeseler, A., & Minh, B. Q. (2015). IQ-TREE: A Fast and Effective Stochastic Algorithm for Estimating Maximum-Likelihood Phylogenies. Molecular Biology and Evolution, 32(1), 268–274. 10.1093/molbev/msu300

Nimmo, C., Millard, J., Faulkner, V., Monteserin, J., Pugh, H., & Johnson, E. O. (2022). Evolution of Mycobacterium tuberculosis drug resistance in the genomic era. Frontiers in Cellular and Infection Microbiology, 12, 954074. 10.3389/fcimb.2022.954074

Noviyani, A., Nopsopon, T., & Pongpirul, K. (2021). Variation of tuberculosis prevalence across diagnostic approaches and geographical areas of Indonesia. PLOS ONE, 16(10), e0258809. 10.1371/journal.pone.0258809

Rukminiati, Y., Mesak, F., Lolong, D., & Sudarmono, P. (2024). First Indonesian report of WGS-based MTBC L3 discovery. BMC Research Notes, 17(1), 176. 10.1186/s13104-024-06825-5

Sinulingga, H. E., Sinaga, B. Ym., Siagian, P., & Ashar, T. (2023). Profile and risk factors of pre-XDR-TB and XDR-TB patients in a national reference hospital for Sumatra region of Indonesia. Narra J, 3(3), e407. 10.52225/narra.v3i3.407

Soedarsono, S., Mertaniasih, N. M., Kusmiati, T., Permatasari, A., Ilahi, W. K., & Anggraeni, A. T. (2023). Characteristics of Previous Tuberculosis Treatment History in Patients with Treatment Failure and the Impact on Acquired Drug-Resistant Tuberculosis. Antibiotics (Basel, Switzerland), 12(3), 598. 10.3390/antibiotics12030598

WHO. (2022). Global tuberculosis report 2022. https://www.who.int/publications/i/item/9789240061729

